# Deep Learning Model for Tumor Type Prediction using Targeted Clinical Genomic Sequencing Data

**DOI:** 10.1101/2023.09.08.23295131

**Authors:** Madison Darmofal, Shalabh Suman, Gurnit Atwal, Jie-Fu Chen, Jason C. Chang, Michael Toomey, Efsevia Vakiani, Anna M Varghese, Anoop Balakrishnan Rema, Aijazuddin Syed, Nikolaus Schultz, Michael Berger, Quaid Morris

## Abstract

Tumor type guides clinical treatment decisions in cancer, but histology-based diagnosis remains challenging. Genomic alterations are highly diagnostic of tumor type, and tumor type classifiers trained on genomic features have been explored, but the most accurate methods are not clinically feasible, relying on features derived from whole genome sequencing (WGS), or predicting across limited cancer types. We use genomic features from a dataset of 39,787 solid tumors sequenced using a clinical targeted cancer gene panel to develop Genome-Derived-Diagnosis Ensemble (GDD-ENS): a hyperparameter ensemble for classifying tumor type using deep neural networks. GDD-ENS achieves 93% accuracy for high-confidence predictions across 38 cancer types, rivalling performance of WGS-based methods. GDD-ENS can also guide diagnoses on rare type and cancers of unknown primary, and incorporate patient-specific clinical information for improved predictions. Overall, integrating GDD-ENS into prospective clinical sequencing workflows has enabled clinically-relevant tumor type predictions to guide treatment decisions in real time.

## INTRODUCTION

Knowledge of a patient’s tumor type is crucial for clinical decision making in cancer, guiding therapy choice and clinical trial enrollment. Tumor type diagnosis is typically performed via histological and immunohistochemical analyses, but these techniques may be inconclusive, especially if a patient’s tumor is poorly differentiated, or when distinguishing between independent primary tumors or clonally-related metastases. For cancers of unknown primary (CUP), which account for 3-5% of patients diagnosed in the US today or 60,000 to 100,000 cases per year, the true tissue of origin is unknown and unable to guide treatment, limiting the number of available FDA-approved therapies and resulting in overall poor prognosis^1–3^. Overall, as incorrect tumor type diagnoses can lead to misinformed treatment decisions and poor patient outcomes, there is a clinical need to improve tumor type diagnosis methods, as a form of enhanced decision support for precision oncology^4^.

Large scale, pan-cancer genomic analyses have shown that many genomic alterations and signatures are associated with specific tumor types. Some alterations are largely specific to a single type, e.g., *APC* loss-of-function mutations in colorectal cancer and *TMPRSS2-ERG* fusions in prostate cancer ^5–7^. Other alterations can narrow the search: *TERT* promoter mutations, for example, are predominantly found in glioblastoma, bladder, thyroid and skin cancers^8^. These associations between genomic alterations and tumor type could provide valuable diagnostic information when other indicators of tumor type are inconclusive. As such, clinical genomic profiling of tumors, already commonly used to identify therapeutically targetable alterations and guide the selection of precision oncology treatments, can be leveraged to guide and refine cancer type diagnoses ^9^.

A major limitation of existing genomic-based tumor type classifiers is their lack of clinical feasibility and scope. Currently, the most accurate classifiers rely on whole genome sequencing (WGS) which is not in routine clinical use, mostly due to cost ^10–13^. Even as the associated cost decreases, the infrastructure required for clinical tumor WGS analysis and reporting does not yet exist. As such, many WGS-based classifiers are derived from small datasets across limited cancer types, likely not representative of true cancer type frequencies seen in larger patient populations, and therefore rendering a significant proportion of cancer patients unpredictable within the context of the model^14–16^. The expected performance of these models on excluded cancer types (i.e. those not available for classification by the model) is heretofore undescribed.

Clinical genomic sequencing is often performed using cancer gene sequencing panels, which target specific mutations in genes known to be recurrently altered in different cancer types. Unlike WGS, these can be readily applied at broad scale. Notably, Memorial Sloan Kettering’s MSK-IMPACT panel is an FDA-authorized clinical test, which profiles somatic and germline alterations in over 500 known cancer genes and has sbeen used to equenced more than 75,000 patients to date ^17^. Developing a classifier from MSK-IMPACT data specifically would resolve many limitations related to clinical feasibility, relying on readily accessible clinical genomic data derived from a large patient cohort representative of true cancer type incidences.

As such, our Center previously established a Random Forest classifier to assist in genome-derived diagnosis (GDD-RF) to predict tumor type from MSK-IMPACT data. GDD-RF provides accurate predictions across 22 major cancer types, with only slightly decreased performance from WGS-based models, and has since been prospectively incorporated into a clinical decision-support pipeline ^18^. Since GDD-RF was released, the MSK-IMPACT dataset more than quadrupled in size, potentially enabling improved accuracy on the original 22 types, expansion to previously excluded tumor types, and inclusion of additional genomic features. Additionally, deep-learning architectures have been shown to improve performance on similar tasks but have yet to be explored in the context of cancer gene classifiers^13^. Here, we describe Genome-Derived-Diagnosis Ensemble (GDD-ENS): a deep learning system designed to directly address these challenges, as well as others through specific modifications for enhanced clinical utility.

Our new model now distinguishes 38 tumor types with higher accuracy than GDD-RF. These 38 types represent 97% of solid tumors available for classification within the MSK-IMPACT cohort. Despite only relying on targeted panel sequencing, the prediction accuracy for GDD-ENS equals or exceeds the accuracy of WGS classifiers. GDD-ENS also allows for easy incorporation of additional, patient-specific clinical information, and provides guidance on rare type and CUP diagnoses. Significantly, GDD-ENS has already been implemented prospectively to provide predictions and prediction-specific feature importance values to clinicians in real time.

## RESULTS

### Clinical Cohort and Development of Ensemble Neural-Net Model

To build GDD-ENS, we aggregated a discovery cohort of 42,694 solid tumors profiled by MSK-IMPACT between 2014 and 2020 with adequate sequence coverage and tumor purity for GDD-ENS development (Methods, Fig. 1A). Previously, we had developed a model, GDD-RF on a small cohort and feature set ^18^. Against this baseline, we added newly collected samples, as well as new genomic features that we had previously established were predictive of tumor type ^13^. Improvements to the model, features, and training set were performed sequentially from the baseline GDD-RF model, and each improvement provided substantial performance improvements on the original 22 types (table S1). The larger training set also allowed us to add 16 additional cancer types to the model; doing so reduces the proportion of excluded patients from 15% of the discovery cohort (with 22 types) to 3.1%. These excluded samples represent 45 different ultra-rare cancer subtypes, the majority of which have fewer than 15 high-purity samples per type. After removing the excluded samples, we split the resulting discovery cohort into training and testing sets for model development. To account for the large variability in overall sample size for the 38 types, we upsampled smaller cancer types to include a minimum of 350 examples per type during training. Any samples from patients included in the training set were removed from our testing set, so that the final size of both sets were for 32,816 samples in the training set and 6,971 in the test set. (Fig. 1A-B).

**Fig. 1.**
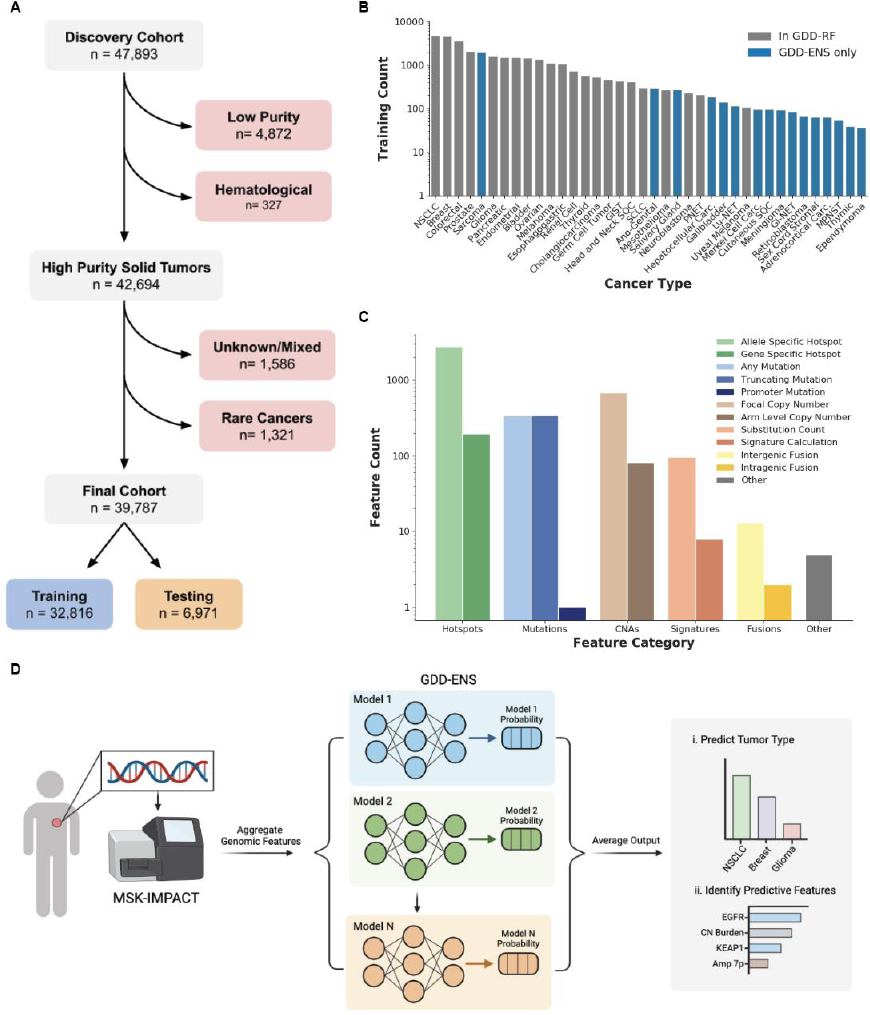
Overview of GDD-ENS model. **(A)** Cohort diagram, detailing samples used to form a training and testing cohort. Hematological refers to blood-based cancers (i.e. Leukemias) sequenced using MSK-IMPACT before development of a separate non-solid tumor assay. **(B)** Training set distribution of cancer types included after expanding GDD-RF to GDD-ENS, colored by model inclusion. Any type with less than 350 examples is upsampled via replacement during training. NSCLC, Non-Small Cell Lung Cancer; GIST, Gastrointestinal Stromal Tumor; SQC, Squamous Cell Carcinoma; SCLC, Small Cell Lung Cancer; PNET, Pancreatic Neuroendocrine Tumor; Lu-NET, Lung Neuroendocrine Tumor; GI-NET, Gastro-intestinal Neuroendocrine Tumor; Carc., Carcinoma; MPNST, Malignant Peripheral Nerve Sheath Tumor. **(C)** Distribution of informative feature types. “Other” refers to clinical features or numerical features representing overall mutational burden across categories. Allele specific hotspot features annotate hotspot mutation (i.e. *KRAS* G12C) whereas gene specific hotspot features only specify the gene altered (*KRAS* Hotspot). CNAs, Copy Number Alterations. **(D)** GDD-ENS workflow from patient to output. All non-clinical features are derived from the MSK-IMPACT sequencing assay, then fed into GDD-ENS. GDD-ENS reports top three tumor type predictions with confidence estimates, along with ten most important features for the top prediction on a patient-specific basis. Workflow overview created with BioRender (https://www.biorender.com/).

Genomic features were derived from MSK-IMPACT data in several broad categories: mutations and indels, focal amplifications and deletions, broad copy number gains and losses, structural rearrangements and fusions, mutational signatures, tumor mutation burden (TMB), microsatellite instability (MSI) score, and sex (Fig. 1C) ^17,19–22^. Feature categories were annotated to varying degrees of specificity; for example we included separate gene-level features indicating presence of any mutation, truncating mutation, or known cancer hotspot mutation for single nucleotide variants and indels, as well as both single-base substitution counts and pre-computed scores for mutational signatures. Our final model includes 4,487 informative features derived from the MSK-IMPACT panel and analysis pipeline (Methods).

The architecture of our final model is a hyperparameter ensemble of ten individual multi-layer perceptrons (MLPs). (Fig. 1D, Methods). We specifically chose this architecture because individual neural net models have been shown to have poorly calibrated confidence estimates and ensembling is a well-established technique to improve calibration while also improving model performance ^23^. The training set was divided into ten training and validation folds, where each validation set represented 10% of the full training set, and each model was trained on the remaining 90%. Models were initialized at the same starting parameters and allowed to optimize to different parameters as a result of their unique training and validation sets. The selected hyper-parameters varied considerably among the models (table S3); often, this diversity improves generalization and out-of-distribution detection ^24^. For each sample, the ten individual MLPs provide a softmax output across all potential tumor types, which is then averaged across all ten models to return a final confidence estimate for each type. The type with the highest confidence after averaging represents the predicted type for the sample.

### Classification Accuracy

We report overall performance of our classifier on the held-out test set from the discovery cohort (N=6,971). We measured performance using a wide variety of metrics, but focused on overall prediction accuracy (proportion of correct predictions across all samples) and macro-precision (class averaged-precision, or the precision of each type averaged, across all 38 types) during optimization and development. The performance of each individual MLP model before ensembling ranged from 73.9 -77.0% accuracy and 57.5– 62.9% macro-precision on the test set (table S4). After ensembling, accuracy was 78.8% and macro-precision was 64.2% (Table 1). Accuracy increases when expanding to the second-highest (87.0%) and third-highest confidence prediction (90.2%) for each sample, as does macro-precision (75.8% and 78.1%, respectively). Moreover, each GDD-ENS prediction is returned with a confidence estimate, representing the model’s predicted probability of correct classification, which we can use to distinguish between high-confidence (≥ 0.75) and low-confidence (<0.75) predictions. In most clinical scenarios we would only consider high-confidence predictions as potentially informative, and low-confidence predictions are less likely to factor into clinical decision-making. The average prediction confidence for all GDD-ENS predictions was 0.84, with 71.9% (5013/6971) of all test samples yielding high-confidence predictions. When restricting to predictions above the high-confidence threshold, accuracy increases to 92.7% and macro-precision to 87.7% (Fig. 2A). Strikingly, GDD-ENS’s high-confidence prediction accuracy and macro-precision are comparable to accuracies reported for WGS-based classifiers, despite being generated from a panel-based dataset (Table 1, Methods) ^10,11,13^. For each method, we also compared the number of predictable types and the in-distribution proportion, or the percentage of discovery cohort samples represented by each specific model’s cancer type labels (Methods). GDD-ENS has the largest number of cancer types and the highest in-distribution proportion, indicating that it can provide relevant predictions for a larger proportion of cancer patients than existing models. GDD-ENS also represents a significant improvement in accuracy, calibration and in-distribution proportion from our originl model (Table 1).

**Fig. 2.**
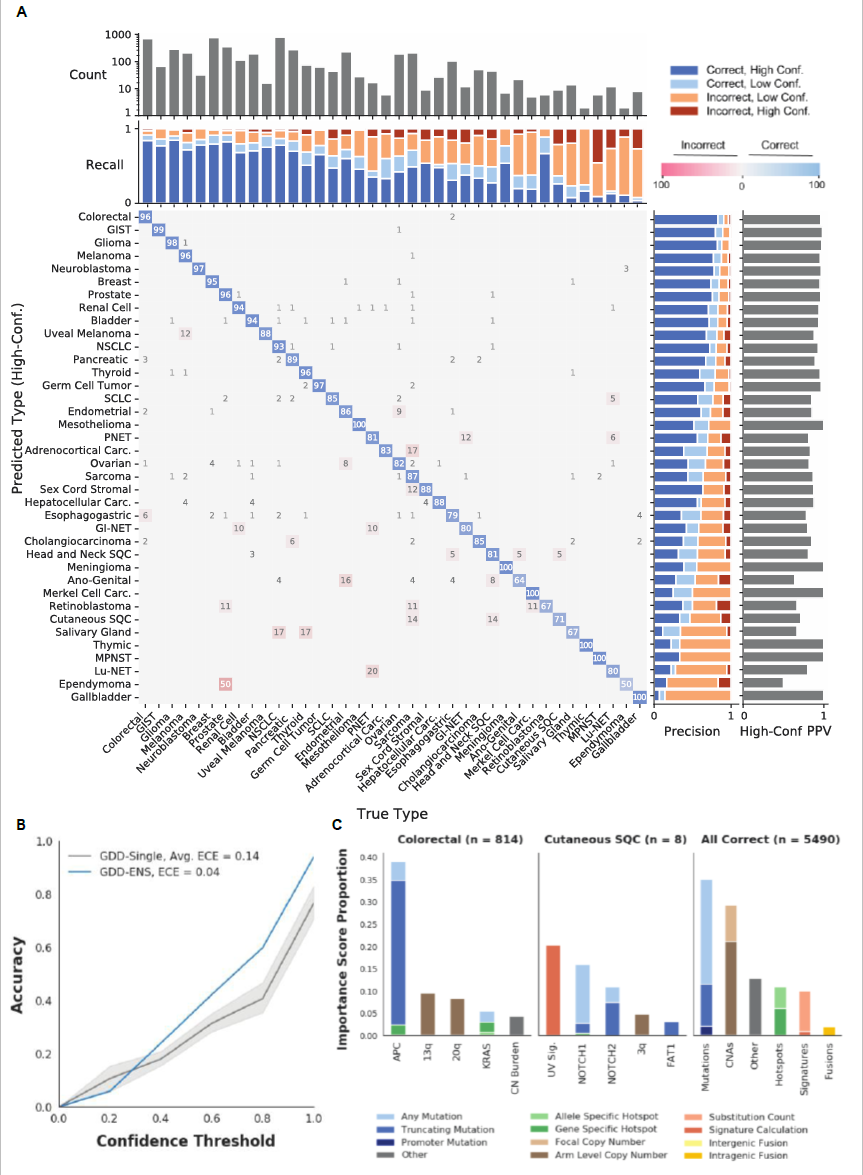
GDD-ENS Performance across cancer types. **(A)** Row-normalized confusion matrix of high-confidence predictions across cancer types. Off diagonal values correspond to proportion of the row, predicted cancer type that is column, true type. The true prostate cancer, predicted ependymoma off diagonal value indicates a single prostate cancer sample that was predicted as ependymoma, likely due to low tumor burden as it just passed the inclusion thresholds for purity. Due to rounding, some rows may sum to more than 100. Upper bar graphs indicate test set count (top) and type-specific recall across confidence levels. Left bargraphs represent high-confidence Positive Predictive Value (High Conf. PPV, right) and type-specific precision. Sorted by overall type precision on both axes. **(B)** Calibration plot for GDD-Single models and GDD-ENS ensemble. X-axis represents maximum confidence after binning outputs into five equally sized ranges, Y-axis represents overall accuracy of all predictions within that range. Blue line represents the GDD-ENS model, dark gray line and shaded regions represent mean GDD-Single accuracy and 95% confidence interval, respectively. Expected Calibration Error (ECE) calculated as per Methods. **(C)** Shapley value score distributions for correct GDD-ENS predictions across individual types (left, middle) and all predictions (right). Importance Score Proportion represents proportion of total Shapley Value scores after summing the absolute Shapley value per feature across the specified subset, normalized as per Methods.

**Table 1.** GDD-ENS Performance and WGS-Based Classifier Comparison. WGS-based methods perform better than panel-based approaches in general, as WGS approaches generate more data and potentially informative features than targeted panels. However, high-confidence GDD-ENS predictions perform similarly or better to most models, on larger set of cancer types that covers a greater percentage of the solid tumor dataset. ID Prop, In-Distribution Proportion, or percentage of high-purity, solid tumor discovery cohort predictable by the classifier’s specific training labels.

| Model | Dataset | Types | Accuracy | Macro-Precision | ID Prop. |
| --- | --- | --- | --- | --- | --- |
| DeepTumour | WGS | 24 | 91% | 91% | 63% |
| CUPLR | WGS | 33 | 89% | 78% | 93% |
| Salvadores-SVM | WGS | 18 | 91% | 86% | 65% |
| GDD-RF | MSK-IMPACT | 22 | 74% | 71% | 85% |
| GDD-ENS | MSK-IMPACT | 38 | 79% | 64% | 97% |
| GDD-ENS<br>High Conf. | MSK-IMPACT | 38 | 93% | 88% | 97% |

We next sought to characterize the classifier accuracy at the level of individual cancer types. Individual type performance ranges from 14.3 – 90.7% before accounting for confidence thresholds (Fig. S2, table S4). The majority of incorrect predictions are low-confidence (75.3%); when only looking at high-confidence predictions, all cancer types have ≥50% precision.

Incorrect high-confidence predictions often reflect tumor biology (Fig. 2A). For example, 10.1% of all high-confidence misclassifications arise between endometrial and ovarian cancer (37/366 predictions). It is well established that high-grade serous or endemetrioid ovarian cancers have very similar genomic characteristics as endometrial cancers, making these cancer types particularly challenging to distinguish ^25^. Despite these errors, GDD-ENS correctly identified at least one endometrial cancer that was initially misannotated as ovarian cancer. Endometrial cancer was predicted with .998 confidence in a patient who was originally diagnosed with ovarian cancer (endometrioid subtype), but a subsequent clonally-related tumor specimen was obtained from the same patient and rediagnosed as endometrial cancer, suggesting that the original diagnosis was incorrect.

Additionally, two tumors annotated as non-uveal melanoma received high-confidence and nearly high-confidence predictions of uveal melanoma (0.91 and 0.74, respectively). This was unexpected, as the genomics of uveal melanoma is notably distinct from other melanomas in that they harbor *GNAQ*/*GNA11* mutations rather than alterations in the *MAPK* pathway more common in cutaneous, acral and mucosal melanoma. However, on further inspection, these two cases both represented the very rare primary CNS melanoma subtype. Previous studies on primary CNS melanomas reported mutation profiles that are more similar to uveal melanomas and are notably distinct from other melanoma subtypes ^26^. Indeed, genomic review of the primary CNS melanoma cases found that both harbored either *GNAQ* or *GNA11* mutations, explaining their GDD-ENS predictions of uveal melanoma.

To ensure that our classifier was applicable across multiple races and ethnicities, we used genetically inferred ancestry as a proxy to assess accuracy across four major ancestry groups ^27^. Despite an overrepresentation of European ancestry within the discovery cohort, the proportion of high-confidence predictions and high-confidence accuracy within each ancestry was not significantly different from the overall high-confidence proportion and accuracy for any ancestry (Fisher’s exact test, p=.05) (Supp. Methods, Fig. S1).

Lastly, we ensure GDD-ENS is well-calibrated by computing the calibration of the full ensemble model and the calibration of each of the individually trained MLPs prior to ensembling. In a well-calibrated model, the confidence value output by the model is approximately equal to the probability that its prediction is correct. Calibration is particularly important when model predictions could be used to guide clinical decisions. Both the calibration plots and the estimated calibration error (ECE) show that GDD-ENS is well-calibrated; and confirm previous reports that ensemble model are better calibrated than individual MLPs ^23^.

### Prediction Specific Feature Importance

To provide insight into the factors driving GDD-ENS’s predictions, we report feature importances using Shapley values ^28^. Shapley values represent the proportion of each output that is contributed by individual features for non-linear systems; their interpretation is similar to proportion of variance explained in regression models (Methods).

We validated this approach for our model by aggregating Shapley values for all correctly predicted instances of each tumor type within the test set, identifying the strongest positive associations per type. Known associations between genomic alterations and tumor types were re-captured through this analysis across all cancer types (Fig. 2C, Fig. S3). We specifically highlight colorectal cancer and cutaneous squamous cell carcinoma as two types with drastically different sample sizes and type-specific accuracies, yet Shapley value analysis identified relevant associations for both cancer types. Colon cancer is one of the most represented cancer types included in our training set, and we find that *APC* truncating mutations are the most important features for colorectal cancer predictions, as expected ^5,6^. Cutaneous squamous cell carcinoma has a smaller sample size with only 8 total correct predictions in our test set, but its top predictive features, UV-Signature and *NOTCH1/2* mutations, are also consistent with the genomics for this cancer type, indicating that GDD-ENS learns important associations even from small numbers of training examples ^29^.

We show the benefit of incorporating driver genes at multiple levels of specificity by highlighting Shapley values for *KRAS*-associated features. *KRAS* is a well-known driver gene present across many cancer types, but the landscape of *KRAS* alterations varies significantly across cancers ^30^. Using strongest positive Shapley values as before, we observe this variation when evaluating correct predictions across pancreatic, NSCLC, esophagogastric and colorectal cancer (Fig. S4). *KRAS* G12C is only associated with NSCLC predictions, while *KRAS* G12D, G12R and G12V are more often implicated in pancreatic cancer, and *KRAS* amplifications are of highest importance to esophagogastric cancer. All 341 genes included in the MSK-IMPACT panel are assessed and included as features in this way, allowing for GDD-ENS to learn and distinguish between specific driver alterations across cancer types, and likely accounting for the increase in performance compared to previous driver based classifiers ^13^.

We then considered Shapley values aggregated across all cancer types, finding that features derived from gene-level mutations are the most informative for tumor type prediction, followed by copy number alterations (**Fig. 2C**). The single most predictive feature, measured by total absolute Shapley value magnitude, was *KIT* mutations in gastrointestinal stromal tumor (GIST), followed by 18p arm level deletion for gastrointestinal neuroendocrine tumor (GNET), and 12p arm level amplification for germ cell tumors, indicating that these associations are highly tumor-type specific. The ability of GDD-ENS to re-capture known trends through Shapley values could enable further exploration of other alteration-tumor type associations with high Shapley values as potential biomarkers for disease or therapeutic targets.

### Performance on excluded cancer types

In clinical applications, it is important to consider all samples, not just those with tumor types represented in the classifier output. GDD-ENS’s 38 cancer types are nearly comprehensive, only 3.1% of samples (n=1,321) in the discovery cohort are from ultra-rare cancer types excluded from the 38 distinct classifiable types. Often it is possible to identify these so-called “out-of-distribution” samples based on GDD-ENS’s output. For example, GDD-ENS predictions on these cases had an average confidence of 63.3%, with only 471/1321 representing high confidence predictions (35.7%) (Fig. 3A), i.e., the majority of the excluded cancer type samples are correctly labeled as low-confidence, thus minimizing their impact in practice. Among the 471 high-confidence predictions assigned to the excluded types, we observed that cases were often predicted as more common cancer types within proper organ systems, likely because most cancers from the same organ system arise from similar molecular pathways (Fig. 3B). For example, over half of all small bowel and appendiceal carcinoma samples were predicted as colorectal. We therefore annotated all high-confidence excluded samples into one of 10 corresponding broad organ systems, and determined how frequently the GDD-ENS predictions aligned with these annotations (Methods). ^31^. For this analysis we removed 205 samples that did not correspond to any of the 10 major organ systems described, resulting in 266 samples with distinct organ system annotations. Overall, 61% of samples were predicted within the expected organ system (162/266). Out of distribution cancers from the gastrointestinal, thoracic, genitourinary and gynecologic systems were consistently predicted within their corresponding systems, indicating that high confidence predictions on rare cancer samples can still guide predictions for the majority of cancer types.

**Fig. 3.**
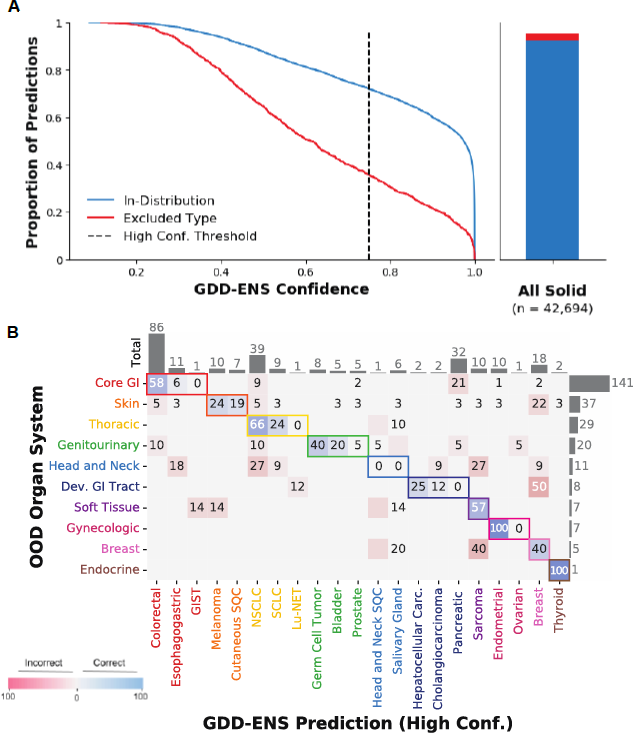
GDD-ENS performance on excluded cancer samples. **(A)** Empirical cumulative distribution function comparing probability of In-Distribution test set and excluded samples (left). 72% of the In-Distribution test set are high confidence, compared to only 36% of excluded samples. Relative fraction of excluded samples among the entire discovery cohort indicates the vast majority of high-purity, solid tumors are In-Distribution (right). HP, High Purity; Conf., Confidence. **(B)** Row-normalized confusion matrix of the organ system of the true type for excluded samples vs the high confidence GDD-ENS prediction organ system. Colors represent broad organ systems annotated, many predictions are conserved within correct organ systems. Rows and columns ordered by total number of excluded type samples within each organ system (right), and number of conserved predictions for each cancer type (top). GDD-ENS predictions correspond to types from the same organ system for 162/266 excluded samples with specific organ system annotations (61%). Dev., Developmental;

### Incorporating additional clinical data into classification

Non-genomic features such as clinical history, histopathological features, and site(s) of metastasis are often available at diagnosis and may further inform cancer type. A natural way to incorporate these features is to compute a new cancer type prior distribution conditioned on these data. For example, to capture information about metastatic site, we computed the cancer type distributions conditioned on each of 19 major metastatic sites using the annotated site of biopsy used for next-generation sequencing. These priors suggest that metastatic site is often informative about tumor type – 55% of skin metastases submitted for genomic profiling originated from primary breast cancers, while 57% of pleural metastatic samples submitted originate from primary non-small cell lung cancer (NSCLC) (Fig. 4A, Fig. S6) ^31^.

**Fig. 4.**
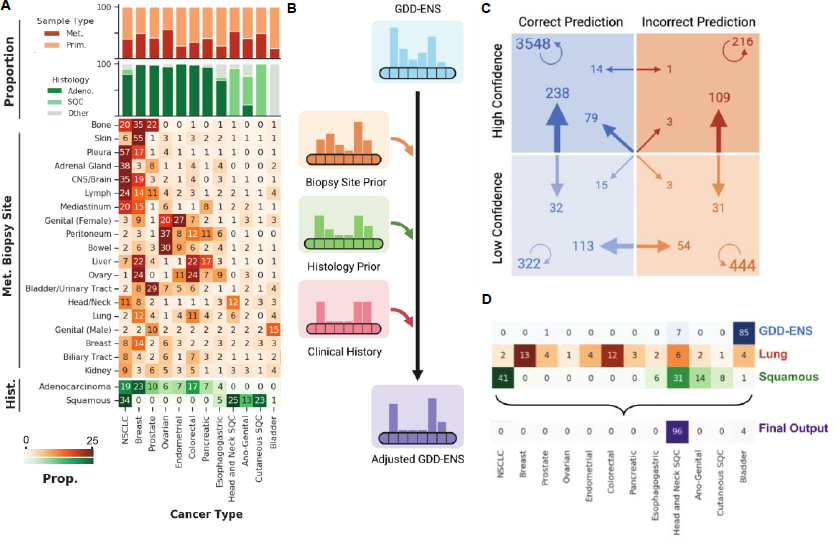
Adaptable prior distribution enables incorporation of non-genomic information for enhanced predictions. **(A)** Proportion of all in-distribution discovery cohort samples that are either primary or metastatic (top), or have broad histological annotations (middle, top) per cancer type. Underlying distributions of metastatic site (middle) and histology (bottom) for all in-distribution discovery cohort samples across 19 metastatic sites and 2 histological subtypes. Heatmaps are row normalized, but only show 10 types (full heatmap in Fig. S6). Met., Metastatic; Prim., Primary; Adeno., Adenocarcinoma; SQC, Squamous Cell Carcinoma;. **(B)** Overview of adaptable prior methodology. **(C)** Flow of results for combination prior using both metastatic site and histology for all test set examples. Arrow base represents pre-adjustment category, arrow head represents post-adjustment. Circle arrows indicates the number of samples that did not change categories after adjustment, i.e. 4618 samples that were correct and high confidence before and after applying the prior. **(D)** Walkthrough of patient with head and neck squamous cell cancer predicted bladder by GDD-ENS with .85 confidence. After applying priors specific to the site of metastasis and annotated histology, the sample was correctly predicted with high-confidence (.96). Overview and flow diagram created with BioRender (https://biorender.com/).

To allow users to incorporate this and other such information on-the-fly, GDD-ENS permits its predictions to be flexibly re-calibrated to account for these features without additional training on non-genomic information. In this “adaptable prior” model, GDD-ENS is augmented with a Naïve Bayes classifier that incorporates the GDD-ENS output and adjusts for any additional non-genomic features, where each feature is represented by its own distribution over cancer types (Methods, Fig. 4B)

We demonstrate the utility of this approach using two broad, non-genomic sources of information: metastatic biopsy site, and tumor histology category. For metastatic biopsy site, we use the 19 major groupings described above to map metastatic biopsy site labels to a broader anatomical site category. We then aggregate all biopsy site-cancer type pairs across all metastatic samples within the training set to compute distributions for the frequency of each cancer type at the biopsy site (Methods). For the histology-based prior, we focus on carcinomas, which represent the majority of our solid tumor dataset, and developed priors based on cancer type distributions within the two major carcinoma subtypes: adenocarcinomas and squamous cell carcinomas. Augmenting genomic predictions with prior information from these broad systems allows for improvements in accuracy and macro-precision when applied to relevant subsets of the test set (Table 2). Furthermore, the frequency of high-confidence, correct predictions increases, and the proportion of incorrect predictions is reduced in both instances. In cases where samples had both histology and metastatic site annotations, we incorporate multiple sources of prior information within our Naïve Bayes classifier approach, which returned similar performance (Table 2, 4C).

**Table 2.**
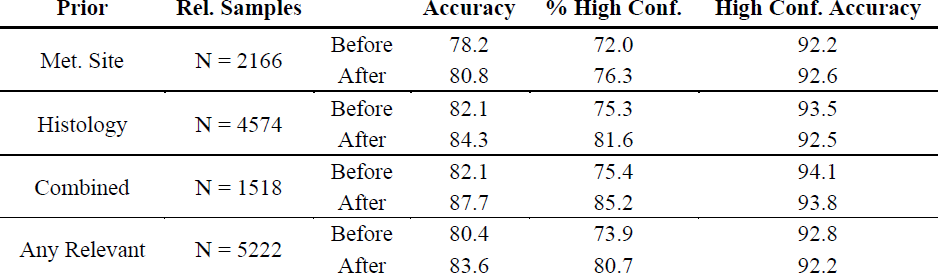
Adaptable Prior Results. Accuracy metrics before and after applying adaptable prior to relevant samples for either metastatic site, histology, or both, where combined represents the samples with both type of annotations, and Any Relevant represents all test samples after applying either Metastatic, Histology, or Combined Priors. Rel. Samples indicates the number of samples within the test set which could potentially be re-calibrated given the samples’ annotations. Overall accuracy and high-confidence proportion increase for all three priors. Rel., Relevent; Conf., Confidence.

We highlight a specific case where applying both adaptable priors was instrumental in producing an accurate prediction. As demonstrated in Figure 4D, we identified a lung metastasis from a patient with head and neck squamous cell carcinoma, initially predicted by GDD-ENS as bladder cancer with .85 probability – a high-confidence, incorrect prediction. After applying both priors, the sample was predicted correctly with .96 confidence. Therefore, applying both available priors not only corrected the prediction, but also increased the prediction confidence enough to surpass the high-confidence threshold for the correct type. Given the success of adjusting outputs given both priors when available, we have made our final adaptable prior extension flexible to multiple sources of prior information, including discrete priors which could be derived from a patient’s specific clinical history.

### Cancer of Unknown Primary Analysis

An important potential application of our method is for cancer of unknown primary (CUP) samples, so that tumor types can be identified through confident genomic-based predictions even when histology is inconclusive. Within the MSK-IMPACT discovery cohort set, there are 1,586 of unknown/mixed cancer types of which 1,441 samples are defined as high-purity CUP samples derived from 1,400 patients. To assess unbiased GDD-ENS performance on CUP samples, we removed any corresponding in-distribution samples from CUP patients within our training set and re-trained GDD-ENS before generating predictions across all the CUP cohort (Methods). While we cannot definitively assess the accuracy of these predictions because most of these tumors remain unclassified by standard histology-based criteria, we identified 26 patients where the CUP sample was followed by a subsequent positively-diagnosed MSK-IMPACT sample performed on the primary tumor, in which the paired samples were found to be clonally related (Methods). GDD-ENS predicted the later confirmed tumor type for 21/26 patients (80.8% accuracy), with the correct indication appearing in the top three predicted types for 24/26 cases. 15/26 samples were predicted with high confidence, all of which represented correct predictions (15/15). These correct predictions spanned 8 distinct cancer types, indicating that GDD-ENS has the capability to accurately guide CUP diagnoses and treatment decisions when histological conclusions are indeterminate for multiple cancers.

This high accuracy encouraged us to expand our analyses to all remaining CUPs, focusing on the high-confidence predictions to quantify model power in the absence of later-confirmed diagnoses. GDD-ENS returned 45.6% of samples with high-confidence, across 36/38 potential cancer types (Fig. 5A). As these cases represent extremely challenging diagnostic scenarios, we further analysed samples with extremely high confidence GDD-ENS predictions; overall, 400 total predictions on CUP samples were >95% confidence, and 255 were >99% confidence, suggesting broad clinical utility of GDD-ENS when applied to cases of indeterminate histopathology-based diagnosis. 906 CUP samples had relevant metastatic site annotations within the 19 previously defined metastatic site labels, 320 had broad histology annotations (e.g., Squamous Cell Carcinoma not otherwise specified) and 190 samples had both, so we applied our adaptable prior distributions to recalibrate CUP predictions. The proportion of high-confidence predictions increased after applying the priors separately (47.1% to 50.8% for metastatic biopsy prior, 42.1% to 54.1% for the histology prior and 42.1% to 58.4% for the combination prior), indicating that adaptable priors for metastatic site and histology can help increase confidence and inform predictions in CUP patients when available.

**Fig. 5.**
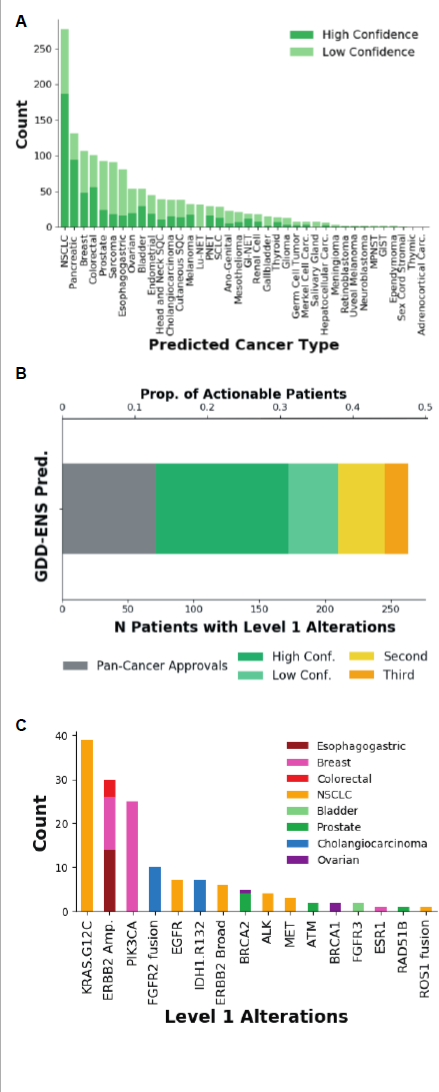
GDD-ENS predictions on CUP patients can identify targetable alterations. **(A)** Distribution of GDD-ENS predictions for all 1,441 CUP patients. **(B)** Barplot indicating total number of patients with targetable alterations after GDD-ENS predictions (bottom axis). Only 550 CUP patients had potentially actionable alterations at level 3B or higher in CUP patients before GDD-ENS predictions, top axis indicates overall proportion of these patients with identified alterations. **(C)** Most frequently identified targetable alteration for only high confidence GDD-ENS predictions. Targetable alteration representing a specific allele change or structural variant are annotated as such, otherwise counts represent combination of broad oncogenic mutations across multiple alteration types and allele changes.

Lastly, a major difficulty in CUP treatment is that most precision oncology therapies are only approved by the US Food and Drug Administration for specific cancer type indications. For example, while a patient with a *ERBB2* amplification and a breast cancer diagnosis would have on-label access to trastuzumab as well as newer antibody-drug conjugates such as T-DM1 and T-Dxd, these potent drugs are not approved for patients with CUP ^32^. We sought to determine if GDD-ENS predictions could increase the number of CUP patients with potentially eligible for available therapies.

We used OncoKB, a precision oncology knowledge base, to annotate the clinical actionability of observed genomic alterations for both CUP and the GDD-ENS predicted cancer type diagnosis, with Level 1 representing an FDA-recognized biomarker predictive of response to an FDA-approved drug within the given cancer type and Level 3B representing a predictive biomarker in another cancer type ^33^. Among the 1,441 CUP patients, only 550 had potentially targetable alterations at level 3B or higher, of which only 71 (12.9% of 550) of CUPs harbored Level 1 alterations – the latter were linked to pan-cancer FDA approvals namely *BRAF* V600E (dabrafenib plus trametinib), *NTRK* fusions (entrectinib or larotrectinib), *RET* fusions (selpercatinib), and microsatellite instability high tumors (pembrolizumab). Thus, 489 CUP patients could potentially benefit from tumor type identification.

After assigning GDD-ENS predictions to these CUP samples, we identified Level 1 actionable alterations in 101 additional patients with high-confidence tumor type predictions, representing a 2.4-fold increase (Figure 5B). The most common targetable alterations identified among these high confidence predictions were *KRAS* G12C mutations (NSCLC), *ERBB2* amplifications (esophagogastric and breast cancer), and *PIK3CA* oncogenic mutations (breast cancer) (Fig. 5C). Notably, an additional 12 CUP tumors with *BRAF* V600E mutations were predicted with high confidence to be colorectal cancer, a diagnosis that would change the approved Level 1 treatment from dabrafenib plus trametinib to encorafenib plus cetuximab ^34^. For those patients in which the top GDD-ENS cancer type did not return any actionable alterations, we expanded our assessment to include lower-confidence predictions, or targetable alterations within the top 3 predicted cancer types (Figure 5B, Methods). With this broadest scope, 263/550 (47.8%) patients with potentially targetable alterations were connected with FDA-recognized treatments, a 3.7-fold increase from the initial 71 patients identified prior to tumor type classification. While GDD-ENS predictions alone are not sufficient to result in the change of a CUP diagnosis to a definitive type for every case, this analysis indicates the potential clinical utility of our classifier in clarifying challenging clinical cases using purely genomic panel sequencing data.

## DISCUSSION

We present a deep-learning tumor type classifier, GDD-ENS, which predicts across 38 distinct cancer types using targeted DNA sequencing data. Specifically, our model is trained using data from the MSK-IMPACT panel, which captures exons from only 341 genes, most of which are common cancer drivers. Despite this, GDD-ENS’s high confidence predictions have performance characteristics comparable to or better than state-of-the-art classifiers trained on WGS data. Unlike previous studies indicating that tumor-type classifiers built on driver mutations alone have poor performance ^13^; we show that with sufficient training dataset size, targeted gene panels can support highly-accurate tumor type classification. Our model features are also well designed to support a panel-based classifier. Building from our prior model ^18^ we capture SNVs, indels and copy number alterations at multiple levels of specificity (i.e. hotspot and allele-specific features for each gene). Incorporating driver gene information in this way allows for the broad spectrum of alterations within a single driver gene to be represented in our feature set for GDD-ENS to use to distinguish similar mutations across diverse cancer types.

Most of GDD-ENS predictions are high confidence (71.9%), and these predictions are reported across a much expanded set of distinct cancer types (Table 1). Expanding to include more cancer types results in some individual types having poor performance when observing all predictions, however we find that simply thresholding outputs based on confidence removes the majority of incorrect predictions, as high-confidence precision values for all cancer types are ≥50%, and 6 types reach 100% (overall average 88%). As these types in some cases represent broad cancer labels of multiple distinct subtypes, future work to develop subtype-specific classifiers should be explored. It should also be noted that our reported accuracy estimate is likely a lower bound. Some predictions that are discordant from the reported, true diagnosis labels could be correct predictions that are misdiagnosed or mislabeled within our test set. While these labelling and diagnosis errors are likely infrequent, GDD-ENS can be used to easily identify these cases for further review, which would also improve overall classifier performance.

In developing a tumor-type classifier specifically for clinical deployment and use, it is important that the model is comprehensive, in that it is able to provide tumor type predictions across the majority of cancer patients. The 38 cancer types recognized by GDD-ENS cover 96.9% of patients within our real-world cohort, representing the vast majority of samples. We also describe expected performance on any rare excluded types, in order to enhance interpretation of the model’s result in a clinical setting. The majority of the 3.1% patients with excluded cancer types are assigned low confidence predictions by GDD-ENS, or have predictions within the correct organ systems, indicating that GDD-ENS can guide diagnoses even on untrained, rare cancer types. Future work could be directed to develop methods that could enable auxiliary rare type classification and detection even with small sample sizes; specifically using few-shot learning for even rarer subtypes with no related cancer types in the in-distribution setting, as these are more likely to have distinct or unrecognizable genomics ^35^.

We demonstrate the utility of an adaptable prior distribution function that enables prediction-specific recalibration and show its potential for improving model performance in multiple contexts. This framework will enable clinicians to incorporate clinical information such as site of biopsy and patient history into tumor type predictions without training the classifier on this information, and could easily be expanded to other contexts. For example, RNA expression or DNA methylation profiles are often strong indicators of cell state and tissue/cell of origin, yet these assays are not as routinely performed in the clinical setting. The adaptable prior offers a way to incorporate data from these assays when available, potentially supplementing an independent, genomic-based classification. While models that are purely trained on these types of inputs, or from auxiliary data such as pathology or radiology slides might be more informative, they also have certain weaknesses compared to a purely genomic classifier as in GDD-ENS ^36–38^. Not only are most expression and methylation-based approaches built on smaller datasets across fewer cancer types, but they can also be confounded by normal tissue contamination and are derived mostly from primary tumor samples, which may not perform as well in the metastatic context ^39,40^. In comparison, GDD-ENS includes primary and metastatic samples in both training and testing, therefore providing an accurate and clinically-relevant estimate of performance, across more cancer types.

The nature of the MSK-IMPACT panel would allow for our model to be easily extensible to most other genomic assays. Using information from broader assays such as WES or WGS as input for GDD-ENS would simply require re-factoring gene and copy number based features to represent only those assessable by MSK-IMPACT. In fact, the MSK-IMPACT panel expanded from 341 to 505 genes over the course of classifier development, but we incorporate samples from each version of the assay by limiting our feature set to the original 341 mutations present in the original FDA-authorized panel for MSK-IMPACT. These 341 original genes represent the most prevalent genes for cancer detection and diagnosis; genes added to future panels were not added as features within our set. Future methods to encode features differently depending on whether they were not present or not assayed could be valuable, as it could allow the full capacity of the MSK-IMPACT panel gene set to be employed, as well as broader extensions. Additionally, we could explore using our adaptable prior method to develop custom priors based on presence or absence of genomic features not included in the original set.

Lastly, we provide GDD-ENS predictions across a set of high-purity CUP patients, with 49.4% of patients predicted with high-confidence, and 29.8% above 95% confidence. For a set of 26 sample pairs where patient initial samples were confirmed through later patient samples, GDD-ENS accurately classifies early CUP samples in 21 of these patients, and achieves 100% accuracy for the 15 samples predicted with high-confidence, which would have enabled faster guided treatments should these predictions have been able to confirm diagnoses in real time. Furthermore, we show that GDD-ENS predictions when applied to patients with CUP could expand patients with potentially targetable level 1 alterations, increasing the proportion of actionable patients from 13% to 48% of a potentially actionable subset. The success of GDD-ENS in patients with CUP could potentiate the model for classification of early cancer samples, an open area of research for many early detection models. As such, work to expand GDD-ENS to cell-free DNA sequencing data is currently ongoing ^41^.

In summary, this study demonstrates a highly-accurate tumor type classification model that was designed to specifically improve upon the clinical relevance and applicability of existing, genomic-based tumor type classifiers through use of a large, fixed-panel dataset and deep-learning model. The potential clinical relevance of our model is immediate: GDD-ENS has already been incorporated into MSK-IMPACT’s workflow in real time to provide cancer type predictions and flag cases for deeper diagnostic review and workup. Overall, GDD-ENS is uniquely positioned for real-world, clinical impact and is capable of providing broad and comprehensive decision support for the many cancer diagnostic challenges.

## MATERIALS AND METHODS

### Study Design

The purpose of this study was to develop and evaluate an algorithm for tumor type classification using genomic features derived from fixed-panel genomic sequencing data. To do so, we collected a dataset of 47,938 samples across 43,625 patients profiled by the MSK-IMPACT (Memorial Sloan Kettering-Integrated Mutation Profiling of Actionable Cancer Targets) clinical series. All patients consented to somatic mutation profiling in a CLIA-compliant laboratory, signed a clinical consent form or enrolled on an institutional IRB-approved research protocol (NCT01775072). Samples from rare or non-distinct cancer types, duplicated patient samples, low-purity samples were removed for later analysis; all remaining samples formed a discovery patient cohort of 39,787 samples. We used data from this cohort to train, test and validate our model, generating 4,487 features per sample. The classifier is a hyperparameter ensemble of 10 multi-layer perceptrons, which used a Gaussian process for hyperparameter optimization during training separately for each model before averaging model outputs for final classification results. We optimized for overall model accuracy and macro-precision (class-averaged precision), as well as proportion of high confidence samples (≥75% probability) and proportion of out of distribution samples during model development, and developed methods for extended clinical utility by incorporating non-genomic features for output re-calibration and reporting performance on Cancers of Unknown Primary.

### Patients and Samples

The discovery cohort included 80 distinct cancer types, which were determined and recorded in real time, along with primary or metastatic site classification, as part of the clinical workup of each case. Surgical pathology reports for all cases were reviewed by molecular pathology fellows in order to ensure accurate diagnoses of each sample and patient, and annotated using appropriate OncoTree label codes representing their detailed type and subtype ^42^. All cancer types and subtypes observed within this initial discovery cohort are annotated in Data File S1.

For classification purposes, we implemented a purity threshold consistent with the GDD-RF model, in order to ensure all samples had sufficient tumor content. Biopsy samples with low tumor purity or signal would likely have reduced sensitivity for detection of genomic alterations, copy number changes, signatures and fusions. This sensitivity reduction would disproportionately affect cancer types with lower frequencies of the genomic alterations, so all samples for which all variants detected had a somatic mutant allele frequency less than 10% and with copy number alterations with an absolute log ratio less than 0.2 were excluded, removing 4,872 samples of our initial cohort.

We excluded 1,586 Cancer of Unknown Primary samples, as well as any diagnosed as a heme-related subtype to focus on classifiable, solid tumor samples for classification. 22 major cancer types were selected for initial development of GDD-RF, which all had at least 40 independent tumors in the original MSK-2017 training set. Given the performance of the initial classifier and sample counts within the expanded dataset, these 22 types were refined and expanded to a final set of 38 types selected for classifier development. Only 1,321 samples (3.1% of high purity, solid tumor discovery cohort) were excluded as a result of this type exclusion. 37/38 types represented distinct tumor types derived from oncotree annotation and pathology review ^42^. Only Ano-Genital is a combination of two originally distinct types, made up of both cervical cancer and anal squamous cell carcinomas, as these types were found to be indistinguishable genomically during classification likely because development of both types are strongly associated with HPV.

The final discovery cohort was split into a training and testing set in an 80:20 split using simple randomization. We removed all redundant patient samples from patients with more than one sample of the same diagnosed cancer type or repeated patient examples in training from our testing set. Our final training and testing cohort included 32,816 and 6,971 samples, respectively. Patient age, sex and other key demographic characteristics of this cohort is shown in table S2.

### Feature Derivation

GDD-ENS includes 4487 features representing mutations, indels, copy number rearrangements, fusions and signatures at varying degrees of specificity. These features represent informative features detected at least once within the discovery cohort training set, any all zero features were removed. While the majority of features included are retained from GDD-RF and are therefore aggregated in the same way, we detail each feature category below, highlighting changes specific to GDD-ENS. All features included are detailed in Data File S1.

#### Mutations

A binary feature indicating presence or absence of a non-synonymous missense mutation for each of the 341 genes present in the original MSK-IMPACT panel was annotated for each sample in the discovery cohort, as well as an additional binary feature specifying the presence of a truncating mutation in these genes. The overall mutational burden attributed to SNVs and INDELs, separately, was included as a numerical feature.

#### Hotspots

We used a dataset of 194 predefined cancer hotspot mutations to generate binary features for presence or absence of gene-specific hotspots for each of the original 341 MSK-IMPACT panel. These hotspots were aggregated from cancergenehotspots.org ^21,22^. We also included 2,725 binary features for presence or absence of any specific hotspot allele within our predefined hotspot list.

#### Copy Number Alterations

The presence or absence of focal amplifications and deep deletions for each of the 341 genes in the panel were included as binary features. Furthermore, binary features indicating the presence or absence of genomic gains and losses for each chromosome arm were calculated from MSK-IMPACT data. Chromosome arms were defined using genomic coordinates in the GRCh37/hg19 human genome assembly, and considered gained or lost if >50% of the arm was affected by segment gain or loss with an absolute value of ±0.2. Lastly, we included a numerical feature representing the overall mutational burden attributed to copy number alterations, defined as the percentage of the autosomal genome affected by copy number gains or losses, as derived from sample segmented log-ratio data.

#### Structual Variants

There are several intronic regions included in the MSK-IMPACT panel designed to assay genes that are commonly rearranged in cancer for structural variants. We identified 13 intragenic fusions and 2 intergenic fusions for feature inclusion from the literature. In GDD-ENS, we included binary features for presence or absence of these structural variants on a gene-specific level, an update from the feature representation in GDD-RF which reported presence or absence on a fusion-specific level. Focusing on the broad genes involved in common structural variants instead of limiting genes to common fusion partners reduced the sparsity of our dataset.

#### Signatures

The presence of mutational signatures was calculated for each sample with at least ten synonymous or nonsynonymous mutations. Any signature representing more than 40% of mutations was annotated as present.

In updating feature representations for GDD-ENS, we expanded the number of included signatures. We also included the single base substitution counts in numerical form for each of the 96 possible substitutions as a way of expanding differential signature strength across samples beyond a binary representation.

#### Clinical Features

Patient sex is included as a binary feature, as certain cancer types are sex specific. In GDD-ENS, we also include MSI-Sensor score, a measure of MSI-Status, as a numerical value ^19^.

### GDD-ENS Model Development

#### Stepwise Model Evaluation (GDD-RF to GDD-ENS)

The original GDD-RF model is a multi-class, random forest ensemble algorithm. While this model was well-suited for the initial problem and training set, when expanding our dataset to include nearly 5 times as many samples and new cancer types, it became necessary to explore other algorithms and feature sets. In order to justify these adaptations, we performed a series of stepwise experiments (table S1). First, using the original training set of 7,791 samples across 22 cancer types (MSK-2017), we updated model architecture from a random-forest to a single MLP. We then updated the features from GDD-RF’s representation as described in the Feature Derivation section of the methods. Next, we updated the dataset to use the full discovery cohort described (MSK-2020). We then updated the architecture again to a hyperparameter ensemble, then finally updated the types from 22 to 38. Each described adaptation was coupled with an improvement in most accuracy metrics (Table S1)

#### Training Procedure

We first trained a single, fully connected feed-forward neural network, which we called GDD-NN. We use a Bayesian optimization approach to select hyperparameters, selecting across multiple hyperparameters: number of hidden layers, number of neurons per hidden layer, learning rate, dropout rate, and weight decay. The final layer of the network was a softmax output, representing the probability distribution of the 38 tumor types. The type with the greatest softmax probability represented the predicted type, and the maximum probability value represented prediction confidence.

The model was trained using Adam, with a batch size of 32 for 200 epochs. Initial hyperparameters were set to the default Adam parameters, but updated during GDD-NN development to represent relevant initializations and optimization ranges, (table S3). We specifically trained the model using gp-minimize, from the scikit-learn scikit-optimization library (version 0.22, RRID:SCR_002577), which evaluates hyperparameter performance using a gaussian process, using gp_hedge as an acquisition function. Each time overall validation accuracy was improved, the model was saved using the given hyperparameter set. To prevent over-fitting, we implemented an early-stopping mechanism which stopped model development after either 5 model updates or 500 calls to gp-minimize. Final model performance was reported across a held-out test set after model training.

As neural nets are often overconfident when presented with training examples with limited data, we developed our ensemble model GDD-ENS, as these architectures have been shown to effectively reduce calibration error. To do so, we split our training set into ten stratified training and validation sets, and trained ten separate models such that individual hyperparameters were selected across each validation set, using the same process as described for GDD-NN. The softmax layer of each model was averaged across all ten models to provide a final prediction and confidence value. We assessed and compared model calibration for GDD-NN and GDD-ENS using estimated calibration error (ECE). Briefly, this represents the difference in expected confidence and reported accuracy within binned confidence levels, normalized by the number of samples reported within each confidence level.

Each model was implemented and trained in Pytorch 1.4.0 (RRID:SCR_018536). All code was written in Python 3.7.6 (RRID:SCR_001658).

#### Comparison to WGS-Based Classifiers

For each WGS-based model we compared the total number of types included to the total number for GDD-ENS. However, these labels were often inconsistent across classifiers, in that some types were more broadly annotated than others, i.e. ‘Sarcoma’ in GDD-ENS, vs ‘Osteosarcoma’ in other models. These represent different subsets of sarcoma patients; a more specific label, while informative, would be incorrect for any sarcomas that are not osteosarcomas. In order to compare the proportion of cancer patients that are ‘in-distribution’ for each of the classifiers, we calculated In-Distribution Proportion, defined as the proportion of the high-purity, solid tumor samples within the discovery cohort that are classifiable with each model’s distinct classifier label set and annotations. In cases where classifiers did not specify included subtypes within a broad cancer type (i.e. only Sarcoma), we included any within the broadest cancer type labelling.

### Shapley Values

We used the SHAP Python package (version 0.37.0, RRID:SCR_021362) to calculate feature importance values per prediction. The upsampled training set was summarized using the SHAP kmeans function to represent baseline values for each cancer type. We then implemented a Kernel Explainer to generate prediction specific importance values. The Kernel SHAP method is model-agnostic and uses a specially-weighted local linear regression to estimate Shapley values for GDD-ENS, given the background distribution from the summarized training distribution. This method was used to calculate important features per prediction across the test set. We report the absolute Shapley value for each feature, specifying whether the importance value was derived from the presence or absence of each feature, excluding SBS transition counts from final outputs due to their lack of interpretability.

To validate this method, we aggregated all feature importance values for correct test set predictions within each cancer type. We summed absolute Shapley values per feature (whether the feature was present or absent) and normalized each feature by the total sum of absolute Shapley values across the correct predictions within the cancer type. As mutation and copy number based features were represented in varying degrees of granularity (i.e. mutation presence, truncating mutations, amplifications and deletions, and hotspot mutations for a single gene), we further aggregated these features to assess overall importance of each feature to correct predictions within each cancer type, and report the top importance broad features per cancer type (Figure 2C, Fig. S3). In a separate analysis, we also aggregated Shapley values by the broad feature category (i.e. Mutation, Hotspot, Copy Number) to determine which classes of features were most important for correct predictions across types. We also analyzed Shapley values for KRAS-associated features only, across any cancer type which included KRAS in the top ten features from this analysis.

For final implementation, we report top ten absolute Shapley values across interpretable features per model prediction.

### Classification of excluded types

We classified 1,321 high-purity, solid tumor samples from our initial discovery cohort as rare, excluded types due to their small sample size and low likelihood to be genomically distinct. These represent 33 distinct tumor types and 23 subtypes of in-distribution cancer types. Organ system labelling for in-distribution and rare types was performed using Oncotree labelling methods ^42^. Organ system was assigned to one of 10 major organ systems, using the Oncotree label subtypes as per *B. Nguyen et al* ^31^. Analysis of rare types and their corresponding organ system predictions was restricted to these 10 major systems: any samples from cancer types that corresponded to multiple organ systems or organ systems outside of these 10 systems were excluded from organ system analysis.

Additional methods for attempting automated out-of-distribution (i.e. rare, excluded type) separation using ensemble statistics or external techniques are described in Supplemental Methods.

### Adaptable Prior Methodology

The adaptable prior distribution function re-calibrates probabilities from GDD-ENS outputs given prediction-specific prior likelihoods using a variation of a Naïve Bayes classifier. We define *P*(*y*_t_) as the likelihood of cancer type *t* for sample *y*. We want to adjust *P*(*y_t_*) given sample and type-specific genomic (*g_t_*) and non-genomic evidence, such as metastatic site (*m_t_*), histology (*h_t_*) or clinical history (*c_t_*). In other words, find *P_adapt_* (*y_t_*) = *P* (*y_t_*|*X*), where *X* = *g_t_, m_t_, h_t_, c_t_*).

Using Naïve Bayes, this expands to 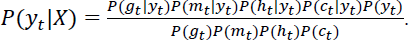

As *P*(*g_t_*|*y_t_*) represents the genomic probability of type *t* for sample *y* as predicted by GDD-ENS, *P*(*g_t_*) is equal to *P*(*y_t_*) for a given type, simplifying the prior to

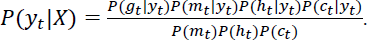

In the case that only a single source of additional clinical information is available to guide predictions all unrelated terms can be similarly simplified, such that the final prior only reflects GDD-ENS probabilities and updated likelihoods given the sample-specific additional information.

We calculate *P*(*y_t_*|*X*) for all *t* to return a final adjustment array which is then normalized, potentially identify a new prediction and confidence estimate for each sample.

We validate adaptable prior using two broad systems: metastatic biopsy site and histology. For the biopsy site adaptable prior, biopsy site labels were mapped to a set of 20 broad organ or organ system labels using ICD Billing codes ^31^. *P*(*x_m_*|*t*) and *P*(*x_m_*) is derived from the metastatic, upsampled training set. For each organ system, we derived site-specific priors given the frequency of each cancer type within the metastatic, upsampled training set seen at that site, using pseudocounts to allow flexibility for unseen (*type, met_site*) pairs. We also include an “Original” prior, for any metastatic site label with too few examples for prior distribution calculation, i.e. a metastatic site label only seen in testing or corresponding to the “Other” system label as per ^31^. Metastatic biopsy site adaptable prior results were then calculated for each metastatic sample within the test set.

Histology site prior was similarly calculated for any samples within the two broad classes of carcinomas: adenocarcinoma, and squamous cell carcinoma. We derived the frequency of each type within each histology using subtype frequencies across the non-duplicated training set, and did not include pseudocounts as prior likelihoods for these are non-flexible. To report the results on test samples, we assume that any sample with a true type within either histology would be observable at the time of diagnosis without requiring a true type indication, so we adjust based on annotated subtype.

When combining multiple priors, i.e. for metastic samples with histology annotated, we multiply across both priors as per Naïve Bayes classification methodology previously described. Patient specific clinical-history priors are not pseudocount adjusted during implementation. Adjusted predictions are calculated similarly regardless of context, multiplying *P(t)* by the adjustment ratio for each type, normalizing, and then reporting highest probability type, and corresponding confidence value.

### CUP analysis

To identify CUP samples with later confirmed diagnoses, we first screened the MSK-IMPACT clinical series with the following criteria: 1) The patient had two or more samples successfully profiled by MSK-IMPACT; 2) one or more of the samples were annotated as CUP; 3) one or more of the samples were not annotated as CUP. Samples of low purity or low quality were excluded as described above. Each patient was manually reviewed by a molecular pathologist (JC or JCC) for clinical and histologic findings as well as corresponding IMPACT sequencing results on the tumors, and the clonal relationship and tumor origin were determined by shared genomic alterations among tumors. Patients who had pathologic diagnoses and primary sites successfully established on the CUP samples from subsequent sequencing results were identified for the analysis. For all CUP predictions, we re-trained GDD-ENS after removing any samples from patients with a CUP diagnosis regardless of the timeline for CUP indication to remove any potential prediction biases from training, using the same training procedure previously described for GDD-ENS. Results for later-confirmed CUP patients are in Data File S1. Adaptable priors were also applied identically for CUP patients.

We used the OncoKB Annotator tool to annotate actionable alterations for all CUP samples ^33^. We first annotated the baseline level alterations for CUP patients, and then re-ran this analysis using the top predicted indication for each CUP patient, before thresholding on confidence. Any patient that was found to have a targetable alteration at baseline was not included in new patient counts when analyzing targetable alterations for CUP after GDD-ENS predictions, although these overlapping cases represent relevant therapeutic regimen changes thus were highlighted separately during analysis. We repeated this process for the second and third highest prediction, similarly removing any patients with an actionable alteration identified from a top prediction when analyzing patient counts after incorporating the second prediction, etc. Gene and alteration frequency was calculated from only the high confidence GDD-ENS predictions.

## Data Availability

All data and code used to produce the presented work are contained within the manuscript, supplement and available online.

https://github.com/mmdarmofal/GDD_ENS

## Acknowledgments

We would like to thank the patients and their families. We would also like to thank Hannes Bretschneider, Chenlian Fu, Dig Vijay Kumar Yarlagadda, Aziz Zafar, Julia Simundza and Peter Zhang for their insights and helpful comments during preparation of this manuscript and codebase.

## Author contributions

Conceptualization: MD, GA, MB, QM

Methodology: GA, EV, MD, MT, MB, QM

Formal analysis: MD, MT

Software: MD, GA, MT, SS, ABR, AS

Validation: SS, MD

Resources: JC, JCC, AV, NS

Data Curation: MD, EV, JC, JCC, AV, MT

Visualization: MD, MT

Writing – Original Draft: MD

Writing – Review & Editing: MD, MB, QM

Supervision: GA, MB, QM, NS

Project Administration: MB, QM

Funding Acquisition: MB, QM

## Data and materials availability

Data and code for generation of full feature tables, as well as code for model training, individual predictions and adaptable prior available at https://github.com/mmdarmofal/GDD_ENS. All other data and full results are present in the paper or the Supplementary Materials and Data File S1.

