## Supplementary Materials for "Deep Learning Model for Tumor Type Prediction using Targeted Clinical Genomic Sequencing Data"

**Supplementary Materials and Methods**

*Comparison of high-confidence accuracy and proportion across ancestries*. We aggregated ancestry labels for each patient as per *Arora* et al (cite). 97.1% of all patients in our training and testing sets had ancestry labels annotated. In order to assess the accuracy within each ancestry, we calculated performance across the test set for each group, focusing our analysis to four ancestries with relevant sample counts and distinct ancestral labels: European (EUR), East-Asian (EAS), African (AFR) and South Asian (SAS). The EUR cohort also included Ashkenazi Jewish patients (ASJ). Admixed (ADM) and Native American (NAM) patients were removed due to indistinct ancestry labelling and sample size, respectively. We also focused on high-confidence predictions, as the cancer type distribution for each ancestry was non-uniform, and GDD-ENS accuracy was highly variable across all cancer types when not implementing a high-confidence threshold. We performed a two-sided Fisher’s exact test on the accuracy for high-confidence predictions within each ancestry, compared to the entire set of high-confidence test set predictions with or without distinct ancestry labels. Our contingency table for this test represented Correct/Incorrect vs Ancestry/Overall for only the high-confidence predictions within the specific ancestry or the test set. We also performed a two-sided Fisher’s exact test for the proportion of high-confidence samples. In this case, the contingency table represented High-Conf/Low-Conf vs. Ancestry/Overall regardless of correctness.

*Auxiliary methods of Out of Distribution detection.* We experimented with several metrics and models to attempt independent separation of in-distribution (ID) from out of distribution (OOD) samples. The first set of methods were generated from GDD-ENS predictions and ensemble statistics. XGBoost classifiers were developed using OOD and ID samples using either only high-confidence OOD samples, or using all GDD-ENS prediction (all_train), but only tested on high-confidence samples as these represented the most relevant cases for identification. Five-fold cross-validation was used, with four folds of train size 7,195 and test size 1,097, and one fold with train size 7,196 and test size 1,096. The following features were each used to build classifiers:

- **probs** (float): the probability assigned to each prediction by GDD-ENS
- **entropy** (float): the epistemic entropy of the averaged probability vector output by GDD-ENS
- **aleatoric entropy** (float): the aleatoric entropy of the averaged probability vector output by GDD-ENS
- **info gain** (float): the information gain of the averaged probability vector output by GDD-ENS, representing the difference between entropy and aleatoric entropy
- **all probs** (vector of floats): the averaged probability vector output by GDD-ENS, with an entry for each GDD-ENS output class
- **logits** **variance** (vector of floats): the variance of the logits for each probability prediction class
- **softmax** **variance** (vector of floats): the variance of the softmax values for each probability prediction class
- **logits** (vector of floats): a vector created concatenating the 10 logits vectors from each ensemble member together
- **softmax** (vector of floats): a vector created concatenating the 10 softmax vectors from each ensemble member together
- **random** (float): a single value drawn from a normal distribution

In addition, the features listed above were combined in to build the following multi-feature classifiers:

- **probs**+**entropy**+**aleatoric entropy**+**softmax variance**
- **probs**+**entropy**+**aleatoric entropy**+**logits variance**
- **probs**+**entropy**+**aleatoric entropy**+**info gain**+**all probs**+**softmax variance**
- **probs**+**entropy**+**aleatoric entropy**+**logits**
- **probs**+**logits**
- **all probs**+**logits**

We also derived a series of classifiers from an augmented version of GDD-ENS which was trained with OOD samples allowing for classification of OOD as a separate “other” cancer type, called GDD-AUG. GDD-AUG was trained identically to GDD-ENS, and OOD was incorporated using 80:20 training splits from the original set of 1,321, high purity OOD samples. GDD-AUG had 76.3% accuracy, and 62% macro-precision, performing slightly worse than GDD-ENS.

OOD classifiers were then constructed using predictions from GDD-AUG. Five-fold cross-validation was used, with four folds of train size 4,073 and test size 1,018, and one fold with train size 4,072 and test size 1,019. In each fold, the number of training ID examples was downsampled to be equal to the number of OOD samples. The following features were each used to build classifiers:

- **aug pred** (int): the class prediction index of the GDD-AUG classifiers
- **aug pred**+**probs**+**entropy**+**aleatoric entropy**

Results for OOD classification are shown in Fig. S5.

**Supplementary Figures**

**Figure S1 | Ancestry Accuracy Differentials**

**(A)** Overall proportion of all annotated ancestries across the training set (left) and testing set (right). Numbers over each bar indicate total within each category. EUR, European; ADM, Admixed; EAS, East Asian; AFR, African; SAS, South Asian; NAM, Native American **(B)** High-confidence proportion (left) and accuracy (right) for European (EUR), East Asian (EAS), African (AFR) and South Asian (SAS) compared to the overall high-confidence proportion and accuracy across the test set. P-values from a two-sided Fisher’s exact test comparing the proportions of these metrics to the overall distribution per ancestry are shown above each bar.

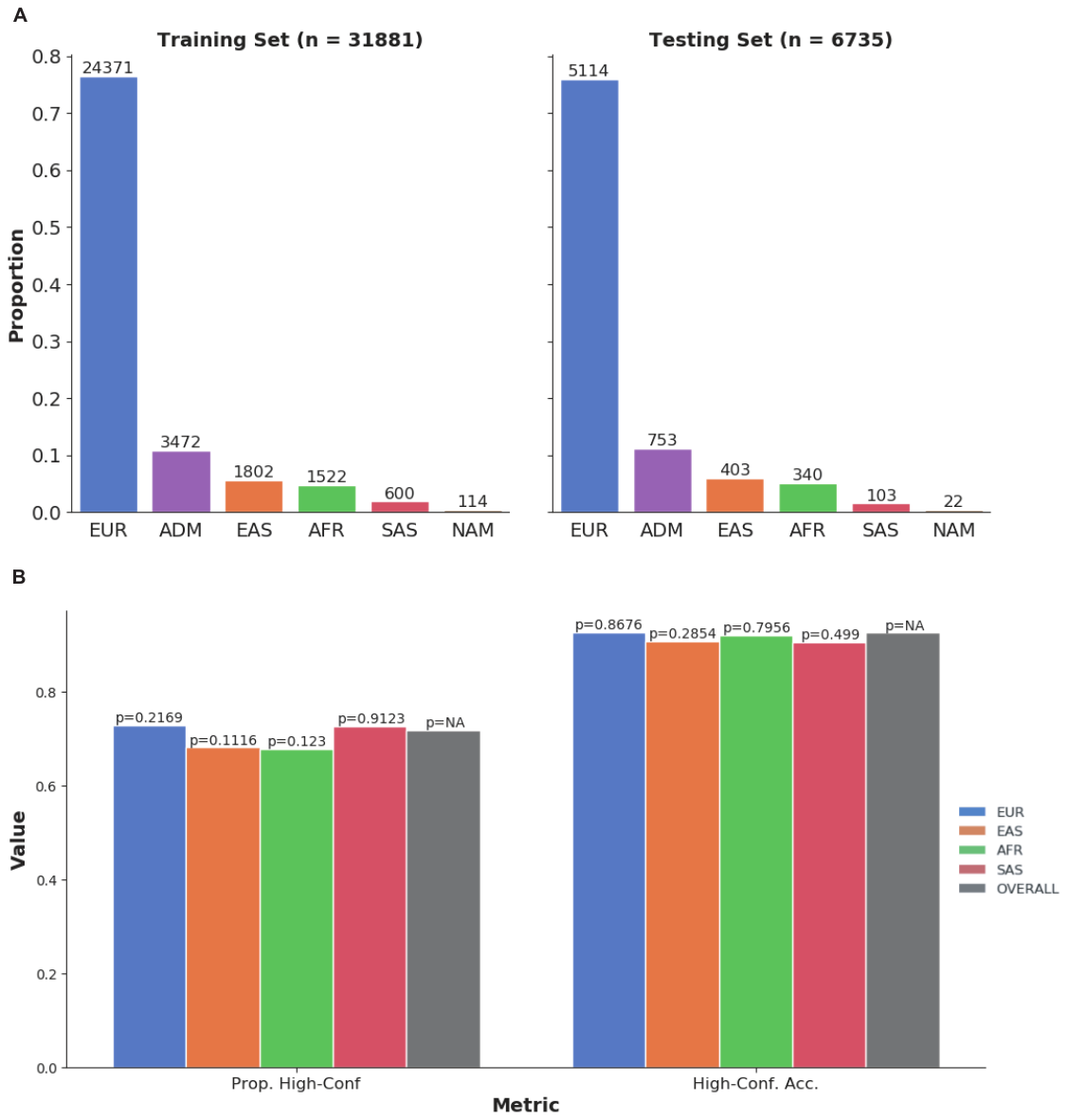

**Figure S2 | Confusion Matrix Across All Confidence Predictions**

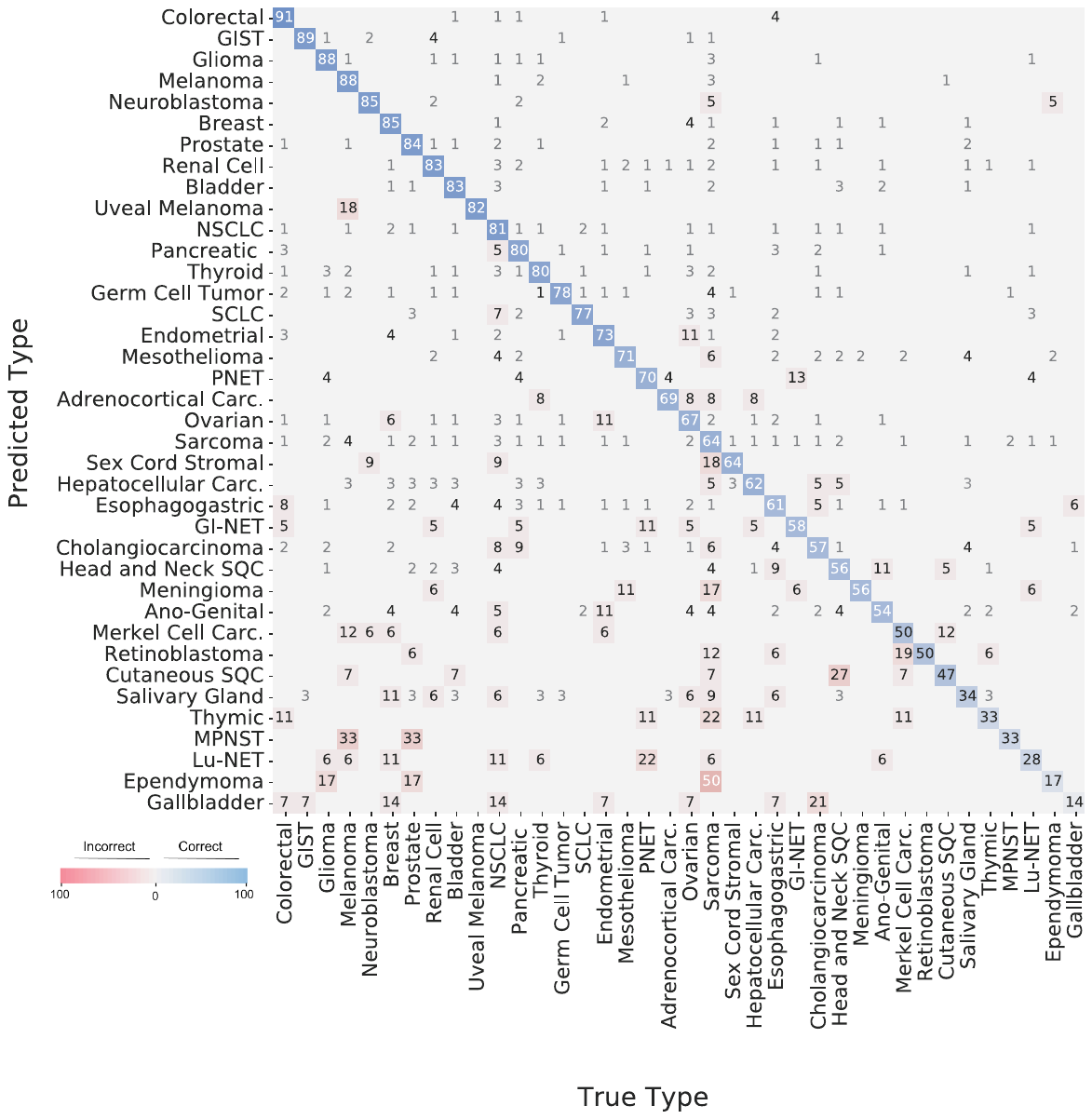
Overall heatmap of top prediction across all confidence values. Heatmap is row-normalized and sorted by overall precision. Off-target values represent proportion of the predicted type, row type that is of the true type along the columns. NSCLC, Non-Small Cell Lung Cancer; GIST, Gastrointestinal Stromal Tumor; SQC, Squamous Cell Carcinoma; SCLC, Small Cell Lung Cancer; PNET, Pancreatic Neuroendocrine Tumor; Lu-NET, Lung Neuroendocrine Tumor; GI-NET, Gastro-intestinal Neuroendocrine Tumor; Carc., Carcinoma; MPNST, Malignant Peripheral Nerve Sheath Tumor.

**Figure S3 | Individual Type Shapley Values**

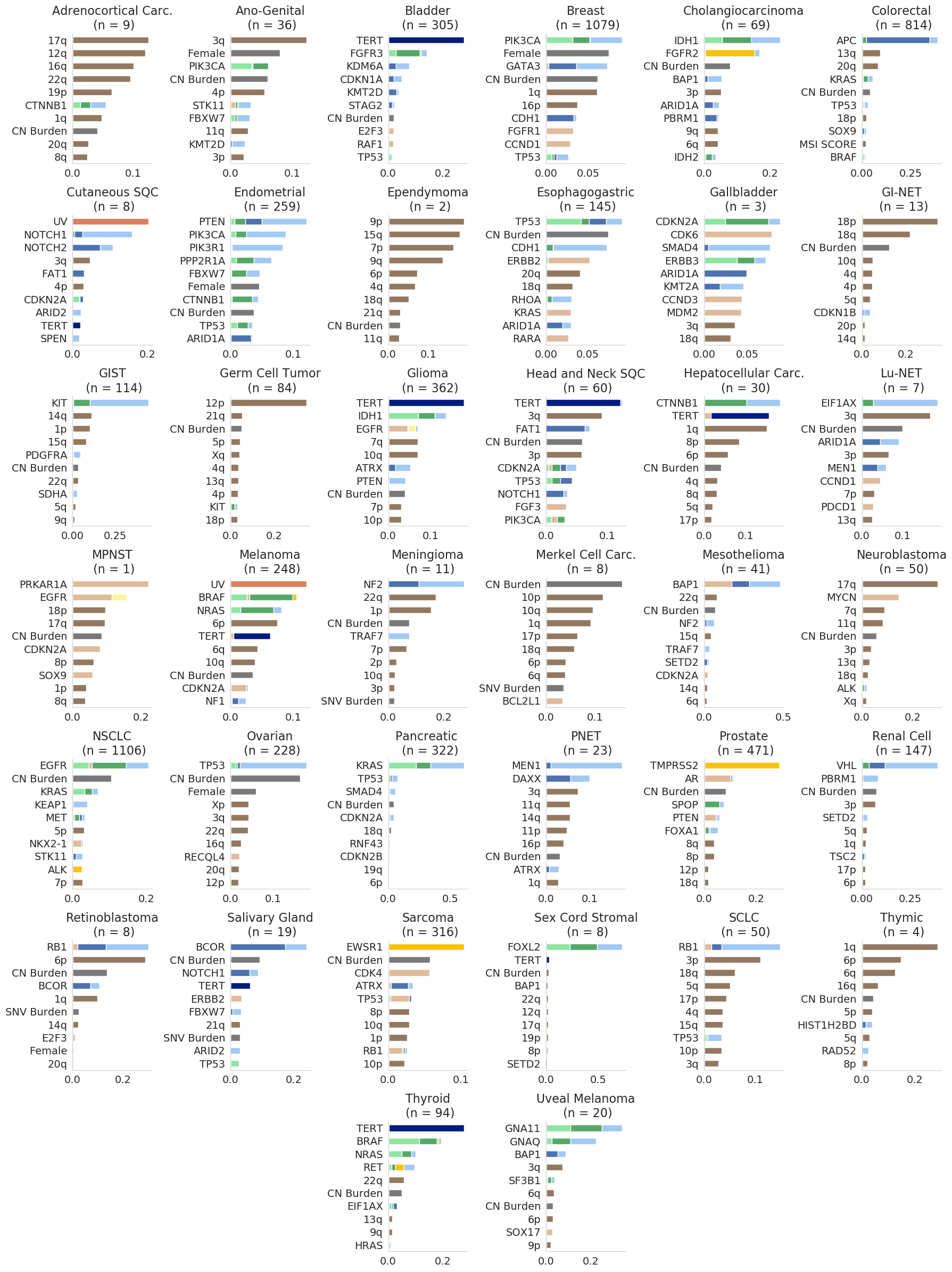
`	Top ten most important features leading to correct predictions of all 38 cancer types included, as approximated by normalized absolute shapley value scores. We sum absolute shapley values per feature, whether the feature was present or absent, and normalized each feature by the total sum of absolute shapley values across the correct predictions within the cancer type. Features that correspond to the same gene or arm segment are grouped, top ten values determined after aggregating across genes or arm segments.

**Figure S4 | KRAS Shapley Values across types**

Normalized absolute Shapley value scores for all KRAS-related features across cancer types with KRAS implicated within the top ten most predictive features per type, based on Fig. S3. KRAS indicate presence of any broad alteration that affects KRAS within a sample, while KRAS hotspot indicates any hotspot alteration, and KRAS Amp reflects a KRAS amplification. All other rows represent features for specific KRAS hotspots. Importance of each specific hotspot differs across cancer types.

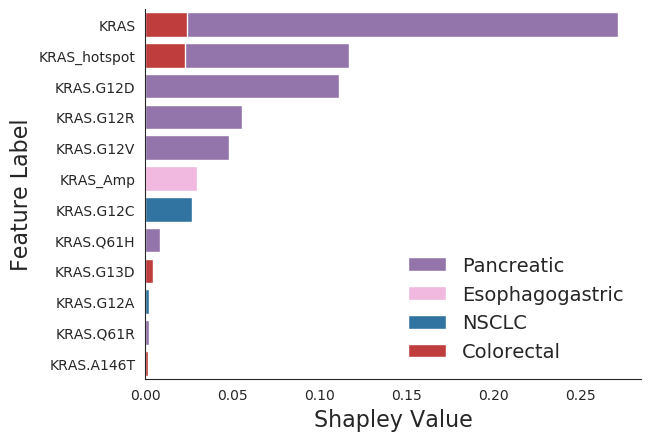

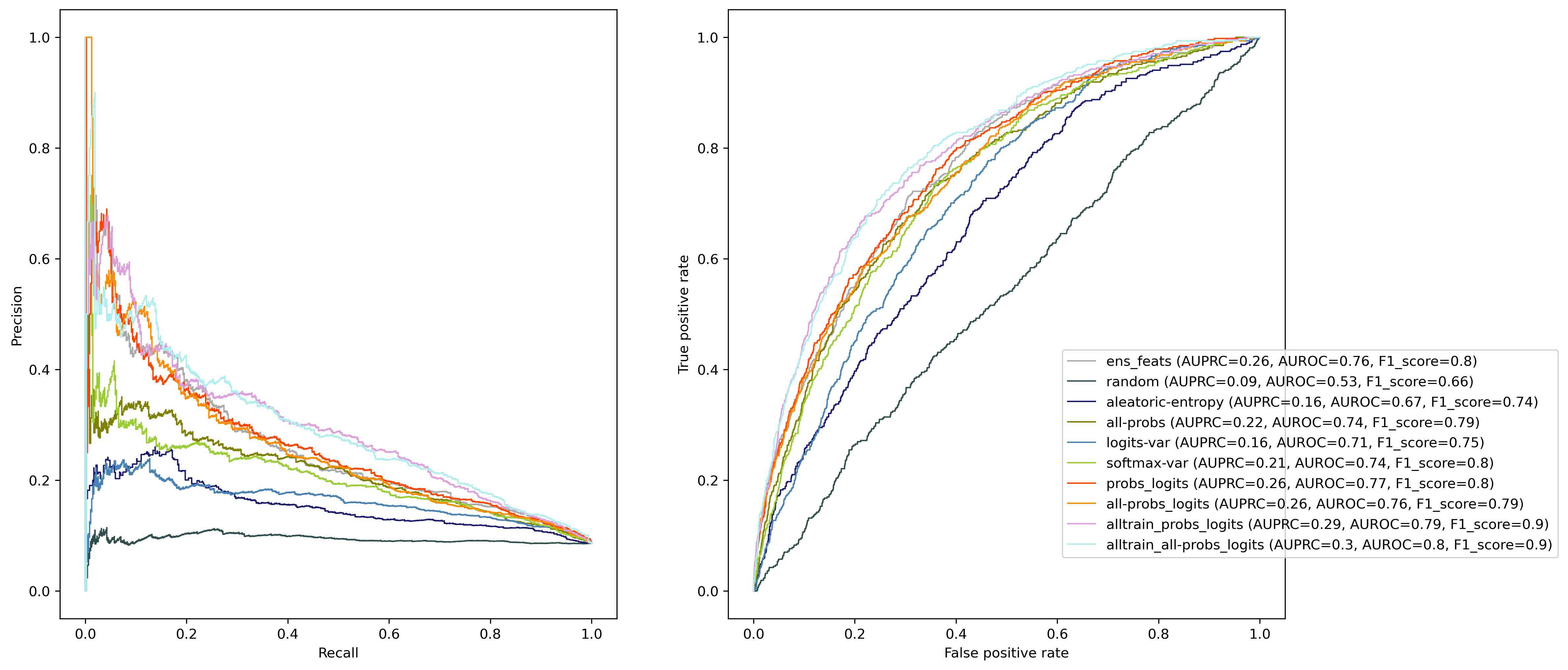
**Figure S5 | OOD detection results**

Precision/Recall (left) and TPR/FPR (right) curves for top classifiers built for OOD detection. Out of all classifiers, the ones that performed best were those built off with the High-Conf. ID and OOD samples using the probs+logits (AUPRC=0.26, AUROC=0.77, F1 score=0.8) and all probs+logits (AUPRC=0.26, AUROC=0.76, F1 score=0.79) features; those built all High-Conf. ID and OOD samples, but tested only on High Conf ID and OOD samples using the probs+logits (AUPRC=0.29, AUROC=0.79, F1 score=0.9) and all probs+logits (AUPRC=0.3, AUROC=0.8, F1 score=0.9) features; and the ensembled logistic regression binary logistic regression classifier (AUPRC=0.26, AUROC=0.76, F1 score=0.8). While these classifiers perform better than random assignment, all methods require further development before potentially applying as a separate classifier for independent OOD detection.

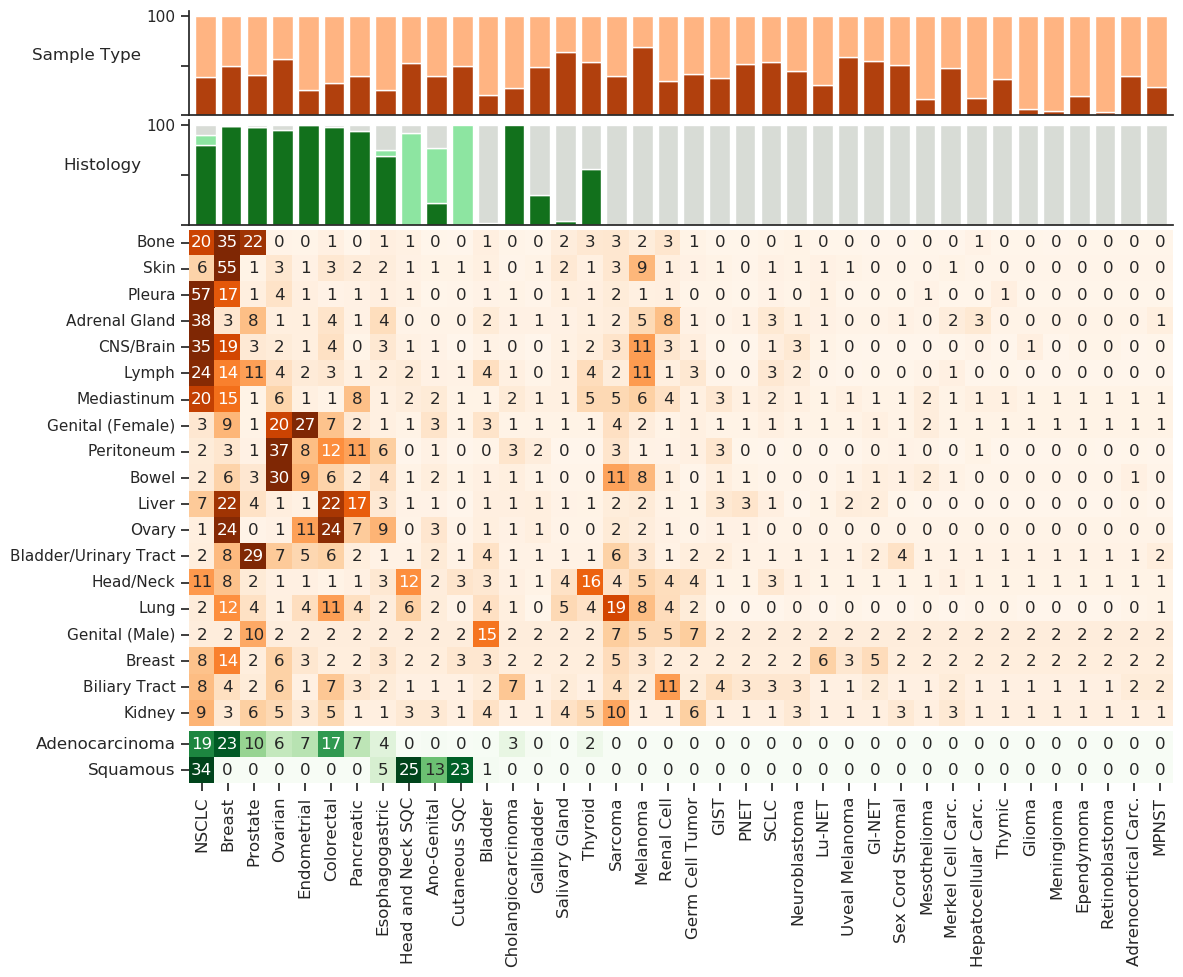
**Figure S6 | Heatmap of Labels Mapped for Adaptable Prior Distributions**

Proportion of all in-distribution discovery cohort samples that are either primary or metastatic (top), or have broad histological annotations (middle, top) per cancer type. Underlying distributions of metastatic site (middle) and histology (bottom) for all in-distribution discovery cohort samples across 19 metastatic sites and 2 histological subtypes. Heatmaps are row normalized, rounding of values means some results do not sum to 100.

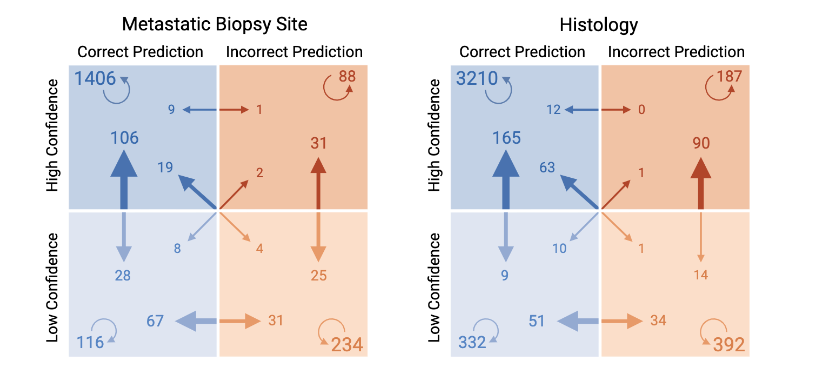
**Figure S7 | Results flow for Met Site, Histology Prior**

Flow of results for combination prior using either metastatic site (left) or histology (right). Metastatic Site prior is only applied to all metastatic samples (n = 2166), histological prior is applied to all test set examples with histological annotations (n = 4571). Arrow base represents pre-adjustment category, arrow head represents post-adjustment. Circle arrow indicates the number of samples that did not change categories after adjustment, i.e. 1406 samples that were correct and high confidence before and after skewing for metastatic biopsy site annotations.

**Supplementary Tables**

***Table S1. Stepwise GDD-ENS model updates and results.***

| **Architecture** | **Feature**  **Set** | **Dataset** | **Types** | **Acc.** | **Macro Prec.** | **% High Conf.** | **High Conf. Acc.** | **High Conf. Macro Prec.** | **% OOD** |
| --- | --- | --- | --- | --- | --- | --- | --- | --- | --- |
| RF | Orig. | MSK-2017 | 22 | 74% | 71% | 62 | 91% | 87% | 15 |
| Single-NN | Orig. | MSK-2017 | 22 | 78% | 76% | 62 | 95% | 92% | 15 |
| Single-NN | Final | MSK-2017 | 22 | 79% | 77% | 64 | 94% | 91% | 15 |
| Single-NN | Final | MSK-2020 | 22 | 82% | 78% | 71 | 95% | 93% | 15 |
| Ensemble-NN | Final | MSK-2020 | 22 | 83% | 80% | 70 | 95% | 94% | 15 |
| Ensemble-NN | Final | MSK-2020 | 38 | 79% | 64% | 72 | 93% | 88% | 3.1 |

MSK-2017: original training set from GDD-RF development (N= 7,791). MSK-2020: updated GDD-ENS discovery cohort training and testing set (N = 39,787) covering samples accrued after initial GDD-RF development. Single-NN, single multi-layer perceptron model; Ensemble-NN, hyperparameter ensemble of 10 multi-layer perceptron models; Acc., accuracy; Conf., confidence; Macro Prec., class-averaged precision. % OOD, out of distribution proportion.

***Table S2 Cohort Demographics.***

|  |  | **MSK-2017 Training Cohort (N = 7791)** | **MSK-2020 Training Cohort (N = 39,787)** |
| --- | --- | --- | --- |
| **Sample Type** | Primary | 4276 | 24249 |
|  | Metastasis | 3515 | 15483 |
|  | Unknown | 0 | 30 |
|  | Local Recurrence | 0 | 30 |
| **Sex** | Female | 3955 | 20355 |
|  | Male | 3810 | 16622 |
| **Age at Sequencing** | mean | 60 | 60.3 |
|  | median | 62 | 63 |
|  | SD | 14.5 | 15.5 |
| **Tumor Purity** | mean | 45.5 | 43.4 |
|  | median | 40 | 40 |
|  | SD | 21.3 | 20.8 |
| **Sequence Coverage** | mean | 718 | 646 |
|  | median | 728 | 641 |
|  | SD | 268 | 212 |
| **Mutations** | mean | 8 | 9.2 |
|  | median | 5 | 5 |
|  | SD | 18.1 | 23 |
| **TMB Score** | mean | 8.4 | 8.4 |
|  | median | 4.9 | 4.4 |
|  | SD | 18.3 | 20.8 |

TMB: Tumor Mutational Burden

***Table S3 GDD-Single and GDD-ENS model parameters and results***

|  | **# FC layers** | **# FC units** | **Dropout Rate** | **Learning Rate** | **Weight Decay** | **Validation Accuracy** | **Test Accuracy** | **ECE** |
| --- | --- | --- | --- | --- | --- | --- | --- | --- |
| **range** | (0, 4) | (5, 2048) | (1e-6, .5) | (1e-5, 2e-3) | (1e-5, 2e-3) | NA | NA | NA |
| **prior** | uniform | uniform | log-uniform | log-uniform | log-uniform | NA | NA | NA |
| **Model 1** | 1 | 1376 | 0.5 | 0.000146 | 0.000787 | 81.27 | 77 | 0.0599 |
| **Model 2** | 2 | 1051 | 0.5 | 0.000222 | 0.000048 | 82.51 | 75.91 | 0.1520 |
| **Model 3** | 2 | 1265 | 0.5 | 0.000145 | 0.000051 | 82.68 | 75.99 | 0.1704 |
| **Model 4** | 1 | 1842 | 0.013185 | 0.000157 | 0.00029 | 81.68 | 75.28 | 0.1287 |
| **Model 5** | 3 | 2048 | 0.000027 | 0.000256 | 0.00001 | 80.3 | 75.4 | 0.1665 |
| **Model 6** | 3 | 1569 | 0.010714 | 0.000269 | 0.000376 | 81.24 | 73.93 | 0.1657 |
| **Model 7** | 3 | 1711 | 0.5 | 0.000084 | 0.00001 | 81.54 | 75.08 | 0.1720 |
| **Model 8** | 0 | 1171 | 0.5 | 0.00001 | 0.000138 | 81.65 | 75.43 | 0.0682 |
| **Model 9** | 3 | 1817 | 0.5 | 0.000123 | 0.000093 | 82.49 | 75.9 | 0.1643 |
| **Model 10** | 1 | 2035 | 0.5 | 0.000182 | 0.000013 | 82.11 | 75.57 | 0.1764 |
| **GDD-ENS** | NA | NA | NA | NA | NA | NA | 78.75 | 0.0499 |

FC: Fully Connected; ECE: Estimated Calibration Error;

| **Cancer Type** | **Train Cases** | **Test Cases** | **Precision** | **Recall** | **High Conf. Precision** | **High Conf. Recall** |
| --- | --- | --- | --- | --- | --- | --- |
| Non Small Cell Lung Cancer | 4893 | 992 | 80.8 | 86.8 | 93.3 | 97 |
| Breast Cancer | 4721 | 942 | 84.8 | 91 | 95.3 | 97.7 |
| Colorectal Cancer | 3536 | 806 | 90.7 | 91.6 | 96.3 | 96.7 |
| Prostate Cancer | 2073 | 417 | 84.3 | 91.4 | 96.3 | 97.2 |
| Sarcoma NOS | 1953 | 380 | 64 | 71.1 | 86.9 | 88.6 |
| Pancreatic Cancer | 1518 | 373 | 80.2 | 83.3 | 89.3 | 94.8 |
| Ovarian Cancer | 1368 | 345 | 66.8 | 63.5 | 82 | 76.8 |
| Endometrial Cancer | 1501 | 335 | 73.5 | 71.8 | 85.5 | 87 |
| Glioma | 1612 | 332 | 87.9 | 90.1 | 97.6 | 98.2 |
| Melanoma | 1120 | 270 | 87.6 | 81.5 | 95.6 | 92.8 |
| Bladder Cancer | 1468 | 257 | 82.8 | 81.3 | 93.8 | 93.8 |
| Esophagogastric Cancer | 1074 | 225 | 60.9 | 53.3 | 78.7 | 66.7 |
| Renal Cell Cancer | 738 | 160 | 83.1 | 80 | 94 | 96.5 |
| Thyroid Cancer | 582 | 125 | 79.6 | 68.8 | 95.5 | 86.5 |
| Cholangiocarcinoma | 535 | 119 | 57 | 50.4 | 85.1 | 78.4 |
| Head and Neck Squamous Cell Cancer | 412 | 108 | 56.3 | 49.1 | 81.1 | 66.7 |
| Germ Cell Tumor | 472 | 93 | 78.3 | 77.4 | 96.8 | 96.8 |
| Gastrointestinal Stromal Tumor | 424 | 87 | 89.3 | 86.2 | 98.5 | 100 |
| Ano-Genital | 350 | 80 | 53.6 | 37.5 | 64 | 72.7 |
| Small Cell Lung Cancer | 350 | 72 | 76.7 | 63.9 | 85 | 77.3 |
| Mesothelioma | 350 | 59 | 71.2 | 62.7 | 100 | 96.3 |
| Salivary Gland Cancer | 350 | 51 | 34.3 | 23.5 | 66.7 | 28.5 |
| Hepatocellular Carcinoma | 350 | 46 | 61.5 | 52.2 | 88 | 81.5 |
| Neuroblastoma | 350 | 41 | 85.3 | 85.3 | 97 | 100 |
| Pancreatic Neuroendocrine Tumor | 350 | 37 | 69.6 | 43.2 | 81.2 | 76.5 |
| Lung Neuroendocrine Tumor | 350 | 30 | 27.8 | 16.7 | 80 | 33.3 |
| Gallbladder Cancer | 350 | 26 | 14.2 | 7.7 | 100 | 12.5 |
| Gastrointestinal Neuroendocrine Tumor | 350 | 21 | 57.9 | 52.4 | 80 | 66.7 |
| Merkel Cell Carcinoma | 350 | 21 | 50 | 38.1 | 100 | 80 |
| Uveal Melanoma | 350 | 20 | 81.8 | 90 | 88.2 | 93.8 |
| Cutaneous Squamous Cell Carcinoma | 350 | 19 | 46.7 | 36.8 | 71.3 | 55.6 |
| Adrenocortical Carcinoma | 350 | 15 | 69.1 | 60 | 83.3 | 83.3 |
| Meningioma | 350 | 13 | 55.6 | 76.9 | 100 | 100 |
| Sex Cord Stromal Tumor | 350 | 13 | 63.6 | 53.8 | 87.5 | 77.8 |
| Thymic Tumor | 350 | 12 | 33.3 | 25 | 100 | 100 |
| Malignant Peripheral Nerve Sheath Tumor | 350 | 11 | 33.3 | 9.1 | 100 | 16.7 |
| Retinoblastoma | 350 | 9 | 50 | 88.9 | 66.7 | 100 |
| Ependymoma | 350 | 9 | 16.7 | 11.1 | 50 | 50 |

***Table S4 Individual Type Performance (Discovery Cohort)***

Conf.: Confidence;
